# Accuracy of ECG Chest Electrode Placements by Paramedics; an observational study

**DOI:** 10.1101/19001321

**Authors:** Pete Gregory, Stephen Lodge, Tim Kilner, Suzy Paget

## Abstract

**Background:** The use of the 12-lead ECG is common in UK paramedic practice but its value depends upon accurate placement of the ECG-electrodes. Several studies have shown widespread variation in the placement of chest electrodes by other health professionals but no studies have addressed the accuracy of paramedics. The main objective of this study was to ascertain the accuracy of the chest lead placements by registered paramedics.

**Methods:** Registered paramedics who attended the Emergency Services Show in Birmingham in September 2018 were invited to participate in this observational study. Participants were asked to place the chest electrodes on a male model in accordance with their current practice. Correct positioning was determined against the Society for Cardiological Science & Technology’s Clinical Guidelines for recording a standard 12-lead electrocardiogram (2017) with a tolerance of 19mm being deemed acceptable based upon previous studies.

**Results:** 52 eligible participants completed the study. Measurement of electrode placement in the craniocaudal and mediolateral planes showed a high level of inaccuracy with 3/52 (5.8%) participants able to accurately place all chest leads. In leads V_1_ - V_3_, the majority of incorrect placements were related to vertical displacement with most participants able to identify the correct horizontal position. In V_4_, the tendency was to place the lead too low and to the left of the pre-determined position whilst V_5_ tended to be below the expected positioning but in the correct horizontal alignment. There was a less defined pattern of error in V_6_ although vertical displacement was more likely than horizontal displacement.

**Conclusions:** Our study identified a high level of variation in the placement of chest ECG electrodes which could alter the morphology of the ECG. From a patient safety perspective, we would advocate that paramedics leave the chest electrodes in situ to allow hospital staff to assess the accuracy of the placements.

**Key messages:** What is already known on this subject

- The recording of a prehospital ECG has become increasingly common in sophisticated Emergency Medical Services across the world
- The accuracy of precordial ECG electrode placement has been studied with other health professionals and has highlighted varying degrees of accuracy.
- Inaccurate electrode placement can lead to aberrant ECG readings and application of unnecessary treatment or the withholding of indicated treatment

What this study adds

- In this observational cohort study, we found significant variation in the placement of the precordial ECG electrodes by UK registered paramedics
- We recommend that paramedics leave the prehospital ECG electrodes in situ to allow hospital staff to assess the accuracy of the placements.

## Introduction

International guidelines for the management of patients presenting with symptoms suggestive of an acute coronary syndrome (ACS) recommend that a 12-lead ECG be recorded by attending emergency medical service (EMS) personnel prior to hospital conveyance.[1-4] The recording of a prehospital ECG has become increasingly common in sophisticated Prehospital EMS Systems and has been shown to significantly increase the proportion of patients who receive Primary Percutaneous Coronary Intervention (PPCI) within 90 minutes of calling the EMS, and to increase the number of ST-elevation Myocardial Infarction (STEMI) patients who receive fibrinolytics in hospital within 30 min of arrival.[5] Patients who receive a prehospital ECG also exhibit significantly lower hospital and 30-day mortality rates than those who did not with most of the differences attributable to significantly lower rates of mortality in STEMI patients.[5] However, the patient benefit that can be derived from the prehospital recording of a 12-lead ECG is reliant upon the ability of EMS personnel to recognise STEMI, or to have access to telemetry to allow another healthcare professional to make the decision, and to accurately place the ECG electrodes. Studies have investigated the ability of EMS personnel to interpret 12-lead ECG recordings in cases of STEMI,[6-9] but none have explored the ability of EMS personnel to correctly place the electrodes.

Incorrect positioning of precordial electrodes presents a risk to patients as it can lead to morphological changes in the ECG,[10-12] with subsequent misinterpretation. The risks are that a patient may receive potentially harmful therapeutic procedures or may encounter a delay in the administration of, or potentially the withholding of, beneficial therapeutic procedures. Accuracy of precordial electrode placement has been studied with other health professionals and has highlighted varying degrees of accuracy. Rajaganeshan et al.[13] found that the correct position for V_1_ was identified by 90% of cardiac technicians, 49% of nurses, 31% of physicians (excluding cardiologists) and only 16% of cardiologists. This study also saw a frequent malposition of V_5_ and V_6_. Medani et al.[14] found that only 10% of participants (doctors, nurses and cardiac technicians) correctly applied all of the leads with the most common errors being the placement of the V_1_ and V_2_ leads too superiorly, and the V_5_ and V_6_ leads too medially. McCann et al.[15] found clinically significant variability in the identification of standardized precordial electrode positions among senior emergency clinicians. From these studies, we hypothesised that there was likely to be a high level of inaccuracy in the placement of the precordial electrodes by EMS personnel.

## Methods

The primary objective of this prospective observational cohort study was to identify the accuracy of precordial lead placement by UK registered paramedics. Participants were recruited at the Emergency Services Show in Birmingham, UK on the 19-20 September 2018. Participants were eligible if they were on the Health and Care Professions Council register (paramedic) at the time of the study, and trained and authorised to record and interpret 12-lead ECGs in the out-of-hospital setting. Recruitment was through posters displayed at the show, promotion by the College of Paramedics (UK professional body) at their seminar sessions, and through word of mouth at the show. Participants were provided with an information sheet and a briefing from the researcher, with an opportunity to ask questions. Written informed consent was obtained from all participants before data were collected. Data were anonymised and information on the performance of individual participants was not made available to anybody outside the research team.

Participants provided professional demographic information relating to their length of experience as a paramedic, the recency of their practice, their academic route to qualification (university route or vocational route), whether they had a specialist role, and the time since their last formal training on ECG electrode placement. Information was collected electronically through the Jisc Online Survey tool (https://www.onlinesurveys.ac.uk/) which allocated a unique identifier to each participant and removed the need to collect person identifiable information. Participants were then asked to place the 6 precordial electrodes on to the chest of a human male model in accordance with their current practice. Before measurement, participants were asked to confirm that they were satisfied with their positioning and were offered an opportunity to make an adjustment if they felt it necessary. Following placement of the electrodes, participants were asked to mark the lead placements on a diagram of bony landmarks with the anterior, mid-, and posterior axillary lines marked on for reference. They were also asked to state, in writing, the landmarks that they used when placing the electrodes and the anatomical positions where each electrode should be positioned. This would allow for comparison between underpinning knowledge and physical performance; only the data from the lead placement is reported here.

Prior to participant enrolment, the correct placements had been pre-determined by two paramedics and an advanced clinical practitioner in accordance with the Society for Cardiological Science & Technology’s 2017 Clinical Guidelines for recording a standard 12-lead electrocardiogram.[16] To maximise the accuracy of our lead placement, we followed precisely the guidelines, measured the mid-clavicular point with a tape measure for V4 accuracy, and had confirmation from an advanced clinical practitioner who was not directly involved with the study. We used a transparent overlay sheet to mark the exact position of our electrodes. The overlay was attached to the model using Transpore™ tape and the position of the corners was marked on the model’s chest using a fine marker pen. The corners of the overlay could then be re-located against the marks and, for consistency, the same researchers placed the overlay into position and completed the measurements. The overlay was pre-printed with 5 mm boxes to assist with the visualisation of the measurement. We used Skintact® FS-50C electrodes as they were typical electrodes for ambulance service use and had a centrally placed connector which was used as a consistent measuring point. Deviation from our positioning was recorded in the craniocaudal (vertical) and mediolateral (horizontal) planes with a deviation of 19mm deemed to be within an acceptable tolerance based on previous studies. Data were input into Microsoft® Excel and then plotted on a scatter graph to show dispersal from the centre point of our electrode. SPSS (version 25.0.0.1, SPSS Inc, Chicago, Illinois, USA) was used to undertake a Two-tailed Pearson Bivariate correlation to establish relationships between the placement of leads across vertical and horizontal planes.

## Ethics

Ethics approval was obtained from the University of Wolverhampton Research Ethics Committee.

## Results

52 eligible participants completed the study. 43 (82.7%) declared no specialist role, 7 (13.5%) said they had a specialist primary care role, one had a critical care paramedic role and one had a role as an ambulance service training officer. The majority (62%) had between one and four years of operational experience as a paramedic and over 86% of participants were current in practice at the time of the study. The route to first registration was mainly via higher education as 43 (82.7%) had either a foundation degree, diploma of higher education or BSc/BSc (Hons) in a paramedic subject. The remaining participants had either followed the vocational route to qualification (15.4%) or a Certificate of Higher Education (1.9%). Only four (7.7%) held a higher degree in clinical practice. There was a wide variation in the time since many participants had received training in ECG electrode placement (Table 1) with a range from less than six months to more than five years.

**Table 1:** Time since last formal training on ECG electrode placement

| Time since last formal training | N (%) |
| --- | --- |
| Within last 6 months | 3 (5.8) |
| Between 6 months and 1 year | 10 (19.2) |
| 1 - 2 years | 11 (21.2) |
| 2 - 5 years | 12 (23.1) |
| > 5 years | 16 (30.8) |

The positioning of the ECG electrode was analysed in respect of the vertical (craniocaudal) and horizontal (mediolateral) planes relative to the pre-determined reference position. Table 2 illustrates the number and percentage of correct placements for each lead, and the range of placements for all leads in both planes. Only three participants were able to correctly place all leads.

**Table 2.**
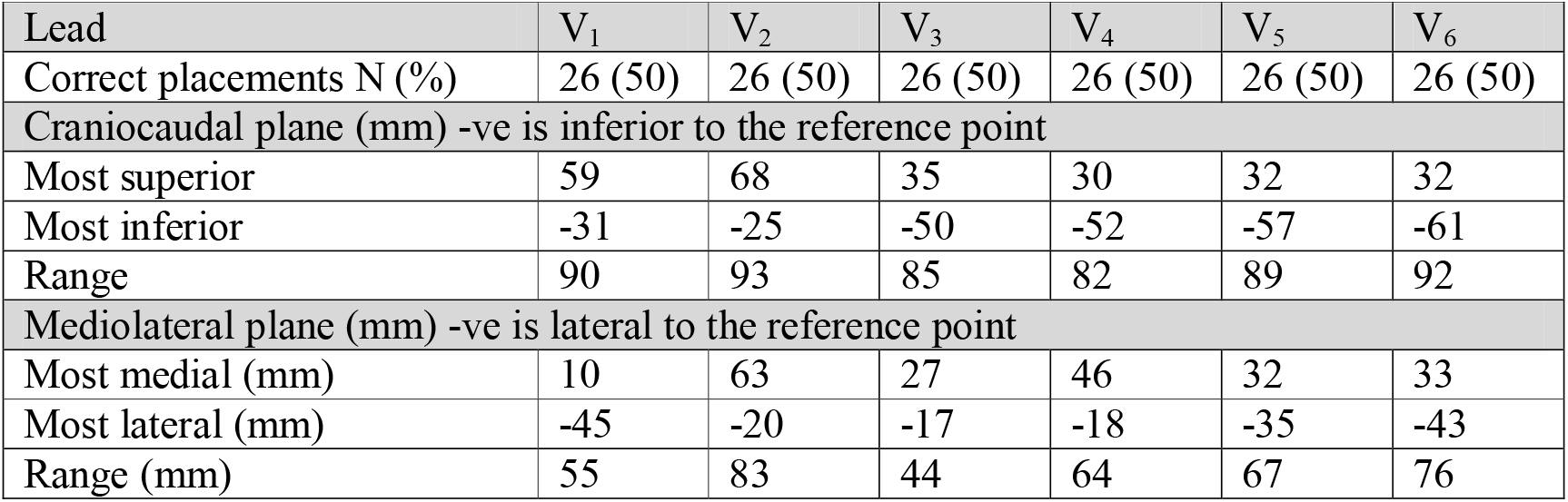
Range of electrode placements (mm) from pre-determined reference point

| Lead | V<sub>1</sub> | V<sub>2</sub> | V<sub>3</sub> | V<sub>4</sub> | V<sub>5</sub> | V<sub>6</sub> |
| --- | --- | --- | --- | --- | --- | --- |
| Correct placements N (%) | 26 (50) | 26 (50) | 26 (50) | 26 (50) | 26 (50) | 26 (50) |
| Craniocaudal plane (mm) -ve is inferior to the reference point |  |  |  |  |  |  |
| Most superior | 59 | 68 | 35 | 30 | 32 | 32 |
| Most inferior | -31 | -25 | -50 | -52 | -57 | -61 |
| Range | 90 | 93 | 85 | 82 | 89 | 92 |
| Mediolateral plane (mm) -ve is lateral to the reference point |  |  |  |  |  |  |
| Most medial (mm) | 10 | 63 | 27 | 46 | 32 | 33 |
| Most lateral (mm) | -45 | -20 | -17 | -18 | -35 | -43 |
| Range (mm) | 55 | 83 | 44 | 64 | 67 | 76 |

The positions of the electrodes are shown in Figure 1. There was substantial variation in the positioning of all electrodes, with patterns of incorrect displacement emerging in leads V_1_ - V_5_. In leads V_1_ and V_2_, the majority of errors were related to the electrodes being positioned too high on the chest. The majority (75% for V_1_ and 67% for V_2_) were able to place the electrode correctly on the horizontal plane. The highest displacement for both V_1_ and V_2_ would have placed the electrode in the second intercostal space.

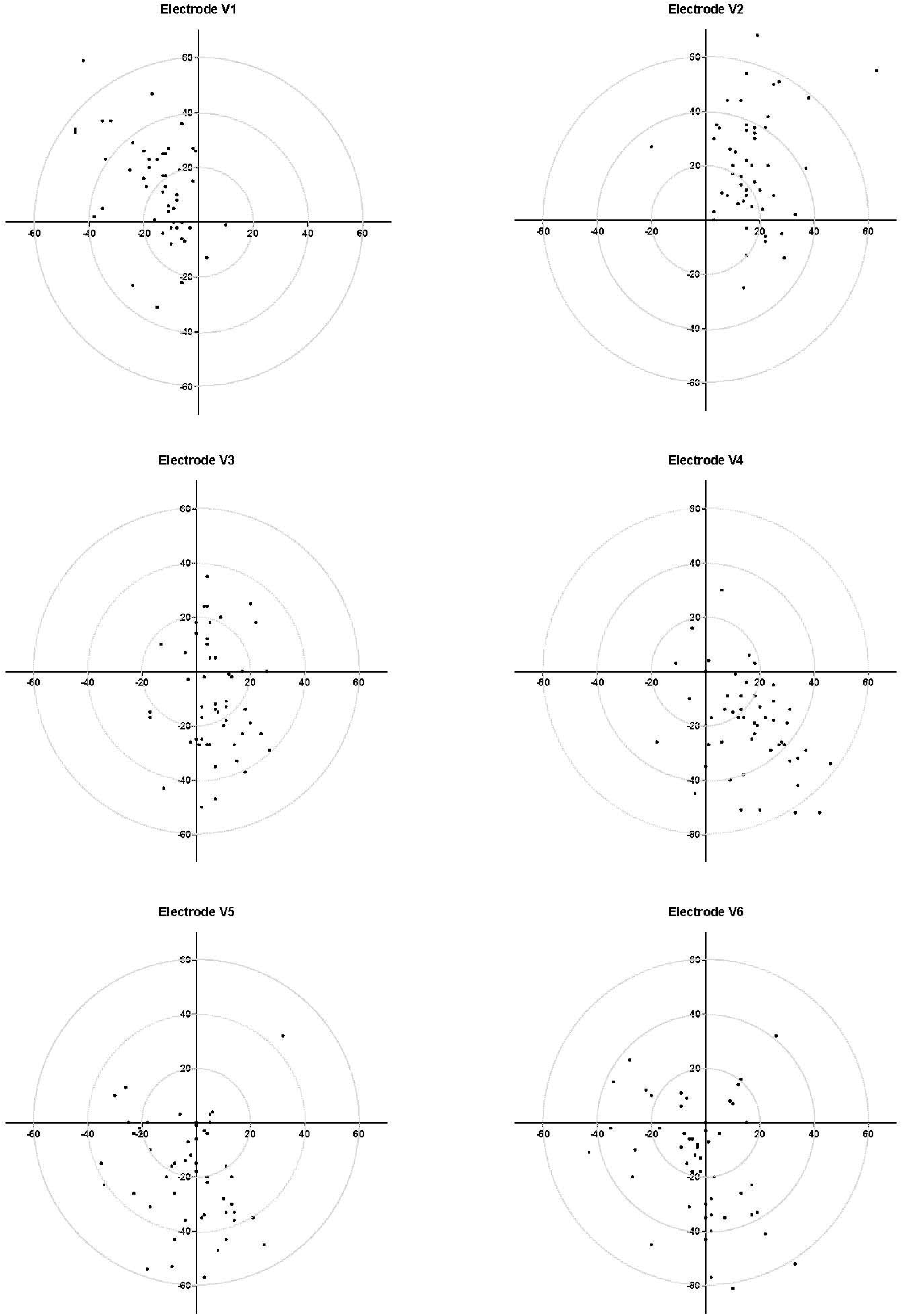

In V_3_, the majority of incorrect placements were related to vertical displacement with most participants (87%) able to identify the correct horizontal position. In V_4_, the tendency was to place the lead too low and to the left of the pre-determined position with only one placement being displaced too high. Placement of V_5_ tended to be below the expected positioning although 77% were able to correctly identify the correct horizontal placement. There was a less defined pattern of error in V_6_ although vertical displacement was more likely than horizontal displacement in terms of absolute numbers and degree of error.

Further analysis of data sought to establish correlation between the placement of leads across vertical and horizontal planes. A Two-tailed Pearson Bivariate correlation was undertaken; these are presented in Table 3.

**Table 3.** Correlation between electrode placements (two-tailed) in craniocaudal and mediolateral planes. Those marked with * were statistically significant at p ≤ 0.05.

| Pearson Bivariate Correlation (Craniocaudal) |  |  |  |  |  |  |
| --- | --- | --- | --- | --- | --- | --- |
|  | V<sub>1</sub> | V<sub>2</sub> | V<sub>3</sub> | V<sub>4</sub> | V<sub>5</sub> | V<sub>6</sub> |
| V<sub>1</sub> |  | .962* | .692* | .348* | .184 | .181 |
| V<sub>2</sub> | .962* |  | .677* | .283* | .203 | .182 |
| V<sub>3</sub> | .692* | .677* |  | .636* | .375* | .295* |
| V<sub>4</sub> | .348* | .283* | .636* |  | .607* | .547* |
| V<sub>5</sub> | .184 | .203 | .375* | .607* |  | .900* |
| V<sub>6</sub> | .181 | .187 | .295* | .547* | .900* |  |
| Pearson Bivariate Correlation (Mediolateral) |  |  |  |  |  |  |
|  | V<sub>1</sub> | V<sub>2</sub> | V<sub>3</sub> | V<sub>4</sub> | V<sub>5</sub> | V<sub>6</sub> |
| V<sub>1</sub> |  | -.288* | -.038 | .049 | .059 | .052 |
| V<sub>2</sub> | -.288* |  | .382* | .263 | .362 | .302* |
| V<sub>3</sub> | -.038 | .382* |  | .486* | .353* | .135 |
| V<sub>4</sub> | .049 | .263 | .486* |  | .705* | .362* |
| V<sub>5</sub> | .059 | .326* | .353* | .705* |  | .832* |
| V<sub>6</sub> | .052 | .302* | .135 | .362* | .832* |  |

## Discussion

In this study, we found significant variation in the placement of the chest electrodes by registered paramedics. Incorrect positioning of electrodes has been well established as a cause of artefact on the ECG, [17– 21] which poses risks to the patient. Patients may receive treatment that is potentially harmful and unnecessary, or they may have appropriate treatment withheld; there is the additional risk of non-conveyance of a patient who should be transported to hospital and the potential danger created by inappropriate transport under emergency conditions. Correct placement of ECG electrodes is also important for reproducibility and diagnosis where serial comparison is undertaken.

Previous studies with other health professionals have identified common misplacement of leads V_1_ and V_2_ [13, 17] with a similar pattern reflected in our study. Placement of both of these leads tended to be significantly higher than the recommended placement with many electrodes situated within the second or third intercostal space. Walsh, [17] has demonstrated that the ECG resulting from such misplacement may generate erroneous patterns such as incomplete right bundle branch block, anterior T wave inversion, septal Q waves, or ST-segment elevation.

The identification of anatomical landmarks is important for the correct placement of electrodes but many participants in our study did not seek to formally identify these landmarks. This meant that when V_1_ was incorrectly placed, V_2_ would be incorrectly placed in a mirror image. The correlation shown between electrodes V_1_ and V_2_ is suggestive that electrode placements were influenced by previous electrode location rather than on identification of anatomical landmarks. For electrodes V_2_, V_3_ and V_4_, it would be expected that a high positive correlation would exist given that V_3_ is positioned midway between V_2_ and V_4_. This was the case in the vertical plane although the relationship between electrodes was not as strong as would have been expected; the reason for this is unclear. As V_2_ was incorrectly placed in a high number of cases in our study, it follows that V_3_ was also misplaced. Electrodes V_4_-V_6_ should be placed at the same horizontal level so again, a high correlation would be expected in the vertical plane. Correlation was strong in these electrodes, but this led to propagation of inaccuracy as misplacement of one electrode influenced misplacement of subsequent electrodes.

We carefully considered our choice of model as other studies have identified obesity and modesty in females as factors linked with poor chest electrode placement. [11, 22] Our chosen model was a male subject of medium build with easily identifiable landmarks so did not present with the complexities of female or obese patients; it is postulated that our results would have revealed greater placement inaccuracy had our model been overweight or female. We do not yet fully understand why there was poor adherence to standard electrode placement in our study, although we did observe that many participants did not locate landmarks prior to placing the electrode. Our second paper (in progress) seeks to understand whether participants knew the correct positions but were unable to translate these to the human body, or whether there is misunderstanding of the correct placement locations.

Our sample size was relatively small, which will impact the generalisability of the results; however, our findings are similar to those from previous studies involving other health professionals and it does suggest a pattern of inaccuracy that causes concern. From a patient safety perspective, we would advocate that paramedics leave the chest electrodes in situ; this will allow hospital clinicians and/or ECG technicians to assess the accuracy of the placement and either utilise the same positioning for a comparative ECG recording, or disregard the findings of the prehospital ECG.

## Conclusion

Our study identified a high level of variation in the placement of chest ECG electrodes by UK registered paramedics. It is not known to what extent, if any, incorrect placement has resulted in incorrect ECG interpretation or patient management but the inaccuracy by our study participants was high and likely to cause morphological changes that could impact on patient treatment. It also raises questions as to the reliability and replication of findings of ECGs from patient to patient and as serial recordings over time for any given patient. We recommend that the prehospital ECG electrodes are left in situ to reduce the risk of unreliable comparative ECG recordings.

Work is needed to understand why this is happening, but emphasis needs to be placed on the importance of identifying landmarks during initial and refresher training.

## Limitations

Our sample size was small and was recruited through a convenience sampling strategy. It is possible that the sample may not be reflective of the wider paramedic population in the UK or internationally but the results do reflect patterns of inaccuracy that have previously been identified in studies involving other health professionals

## Data Availability

The data are deidentified participant data and will be available from Pete Gregory. Data will be available immediately following publication and available for five years. The study protocol, consent forms and participant information sheets will also be made available on request.

